# Low-cost, Smartphone-based Specular Imaging and Automated Analysis of the Corneal Endothelium

**DOI:** 10.1101/2020.12.01.20242271

**Authors:** Sreekar Mantena, Jay Chandra, Eryk Pecyna, Andrew Zhang, Dominic Garrity, Stephan Ong Tone, Srinivas Sastry, Madhu Uddaraju, Ula V. Jurkunas

## Abstract

**Purpose:** Specular and confocal microscopes are important tools to monitor the health of the corneal endothelium (CE), but their high costs significantly limit accessibility in low-resource environments. In our study, we developed and validated a low-cost, fully automated method to quantitatively evaluate the CE using smartphone-based specular microscopy.

**Methods:** A OnePlus 7 Pro smartphone attached to a Topcon SL-D701 slit-lamp was used to image normal the central corneal endothelium using the specular reflection technique. Images were automatically processed on-device and endothelial cell density (ECD), percentage of hexagonal cells (HEX), and coefficient of variation (CV) values were determined using our novel image analysis algorithm. The morphometric parameters generated from the images taken by Tomey EM-4000 specular microscope were compared between the testing modalities.

**Results:** No significant differences in ECD (2799 ± 156 cells/mm^2^ vs 2779 ± 166 cells/mm^2^; p=0.28) and HEX (52 ± 6% vs 53 ± 6%; p=0.50) computed by smartphone-based specular imaging and specular microscope, respectively, were found. A statistically significant difference in CV (34 ± 3% vs 30 ± 3%; p<0.01) was found between the two methods. The concordance achieved between the smartphone-based method and the Tomey specular microscope is very similar to the concordance between two specular microscopes reported in the literature.

**Conclusions:** Smartphone-based specular imaging and automated analysis is a low-cost method to quantitatively evaluate the CE with accuracy comparable to the clinical standard.

**Translational Relevance:** This tool can be used to screen the CE in low-resource regions and reveal the need for further investigation of suspected corneal endotheliopathies.

## INTRODUCTION

The corneal endothelium (CE) is located on the posterior corneal surface and plays a key role in maintaining the cornea in a state of relative dehydration by acting as a selective leaky barrier and an active metabolic pump, which helps to maintain its optical transparency.^1^ The CE is composed of a monolayer of hexagonally shaped cells that rest on a specialized membrane called Descemet’s membrane. Proper size, shape and cell density of the CE are vital to maintaining the corneal hydration balance and are indicative of CE function. The integrity of the CE can be compromised primary corneal endotheliopathies, such as Fuchs endothelial corneal dystrophy (FECD), or secondary endotheliopathies due to intraocular surgery, such as cataract surgery. Data collected from a national managed-care network in the United States found an approximate prevalence of 897 cases of endothelial disease per 100,000 people, 60.4% of which were attributed to FECD.^2^ Furthermore, FECD is the most common cause of corneal transplantation performed in the US, accounting for 68.3% (almost 32,000) of keratoplasties performed annually.^3^ Unmanaged corneal endotheliopathies can lead to corneal edema and vision loss.

The ability to evaluate the morphological characteristics of the CE is critical to diagnosing and monitoring patients with corneal diseases. Currently, specular microscopy (both contact and non-contact) and *in vivo* confocal microscopy are capable of imaging the CE and allow for morphometric analysis.^4^ These commercially available microscopes automatically determine endothelial cell density (ECD), the percentage of cells which are hexagonal (HEX), and coefficient of variation (CV), which are important parameters in evaluating the CE. However, these microscopes have high operation costs ($25,000-$30,000) which limit accessibility in resource-scarce settings. Though this is especially significant for developing countries, even in the United States specular microscopes are prohibitively expensive for many medical practices due to the limited indications for which Medicare provides reimbursement.^5^ Therefore, there is a need for a low-cost and reliable method to image and evaluate the CE.

Smartphone-based specular microscopy has been described as a low-cost technique to image the CE at subcellular resolution.^6^ This method uses a smartphone attached to a slit-lamp ocular and utilizes the specular reflection technique to image the CE. However, this method has not been validated or compared to currently available specular microscopes. Furthermore, prior studies required manual cell annotation of smartphone-based specular images by trained ophthalmologists, making this technique labor-intensive and limiting its ability to be deployed in a clinical setting.^7^

Automated analysis of smartphone-based CE images would make this technique clinically feasible. However, this analysis is a complex image processing task. Uneven illumination across the image, the presence of artifacts, and low resolution are all factors which make segmentation of cell borders and subsequent calculation of ECD, HEX, and CV very challenging.^8^ Images acquired via smartphone-based specular microscopy are even more difficult to analyze than those obtained using a traditional specular microscope, as they have lower resolution and more noise.

Over the past two decades, many researchers have proposed image processing methods to accurately segment cell borders and compute the morphological parameters of endothelial cells. Recent papers have described segmentation methods based on convolutional neural networks (CNNs) which have the potential to be more accurate than existing techniques.^9,10^ However, these methods require a very large training dataset with corresponding ground truths, in which cell borders need to be manually delineated by ophthalmologists. Additionally, these CNNs are very computationally expensive and are difficult to deploy on a mobile device.

Many traditional segmentation approaches are based on the KH-algorithm, which uses directional filtering to identify cell borders.^11^ We built a novel image processing pipeline that combines the KH segmentation algorithm with light normalization, denoising, and artifact removal techniques to enable accurate analysis of smartphone-based specular images. Our platform computes clinically-relevant parameters, such as endothelial cell density (ECD), hexagonality (HEX), and coefficient of variation (CV), and the complete analysis can be run in under 5 seconds on a mobile device, without the need for an internet connection.

In this study, we describe a smartphone-based CE imaging technique and our fully-automated analysis pipeline and validate it against the clinical standard. We compare morphometric analysis parameters from healthy participants as determined by our smartphone-based analysis to those reported by a commercially available non-contact specular microscope.

## MATERIALS AND METHODS

### Data Acquisition

This study was conducted at the Sri Kiran Institute of Ophthalmology, a referral eye hospital in Andhra Pradesh, India, with examination dates between January 2nd, 2020 and January 9th, 2020. This study was approved by the Institutional Review Board of the Sri Kiran Institute of Ophthalmology and was in compliance with the Declaration of Helsinki. Informed written consent was obtained from all participants.

We recruited 15 healthy human subjects without known ocular diseases. All participants enrolled were individuals who had appeared for a routine eye examination.

The central cornea of both eyes of each participant was imaged using the Tomey EM-4000 specular microscope (Tomey Corporation, Arizona, USA). The ECD, HEX, and CV values outputted by the specular microscope were recorded. Subsequently, images were acquired using a smartphone (OnePlus 7 Pro, OnePlus Technology, Shenzhen, China) fastened to one ocular of a Topcon SL-D701 slit-lamp (TopCon Corporation, Tokyo, Japanese) using a smartphone-telescope adapter mount (Gosky Optics, Atlanta, GA), as shown in Figure 1. The specular reflection technique was used to capture the CE at subcellular resolution. The slit-lamp height and width were adjusted to 4 mm and 2 mm, respectively. The light source was positioned 45 degrees relative to the optical axis of the slit-lamp, the ocular was adjusted to 40x zoom, and the backlight was placed on the lowest brightness setting. An ISO of 800 and shutter speed of 1/125 were used to capture images of the CE. The smartphone was manually focused until the characteristic hexagonal pattern of the CE was clearly visible, and the image was captured. A technician performed all imaging, and the total capture and analysis process took approximately 2-4 minutes for both eyes.

**Figure 1:**
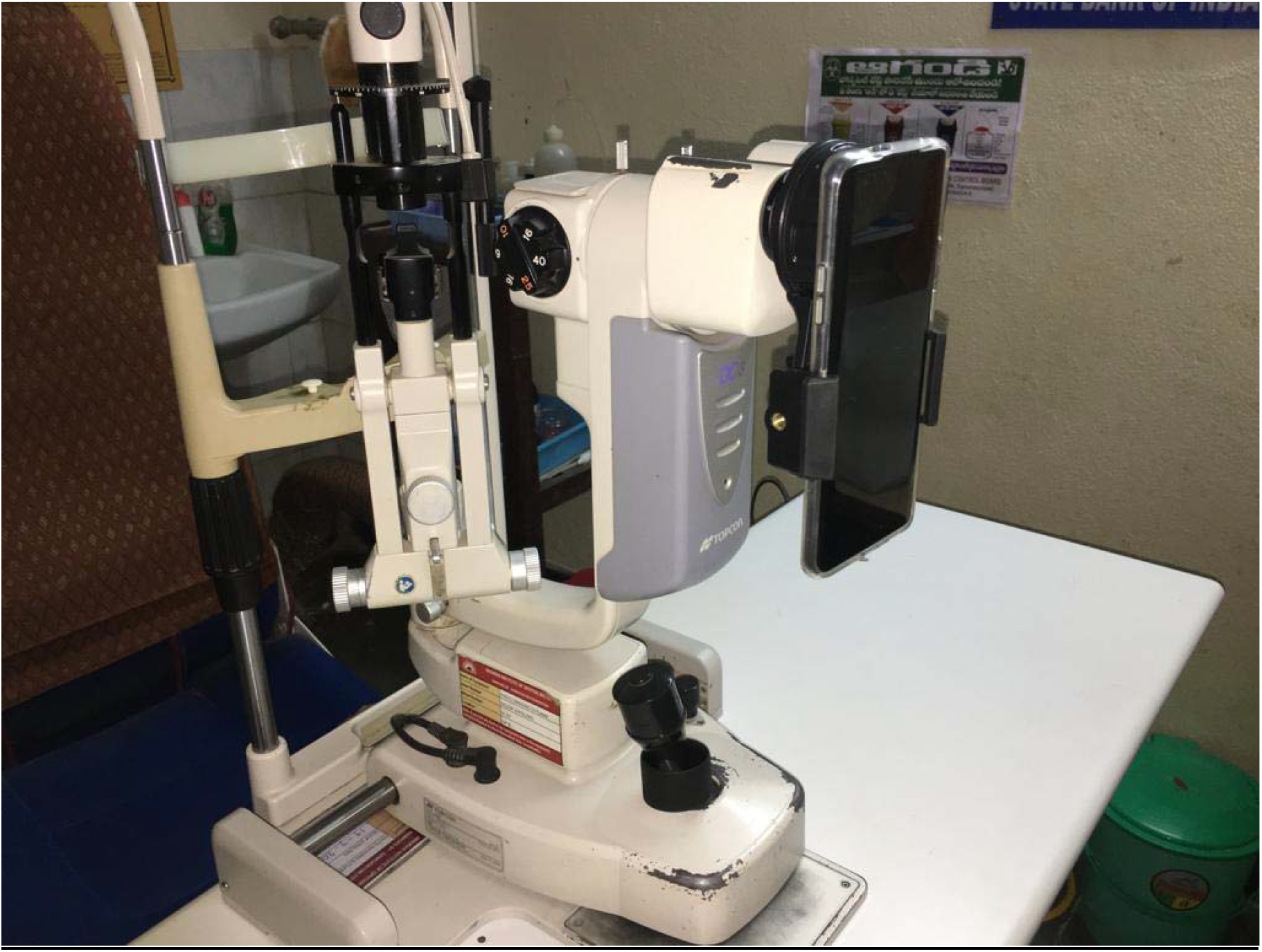
Imaging Setup of Smartphone-based Method. OnePlus 7 Pro smartphone is shown attached to Topcon slit lamp.

An example image acquired using this smartphone-based method and the corresponding image of the same eye captured using the Topcon specular microscope is displayed in Figure 2. The smartphone app user workflow is represented in Figure 3.

**Figure 2:**
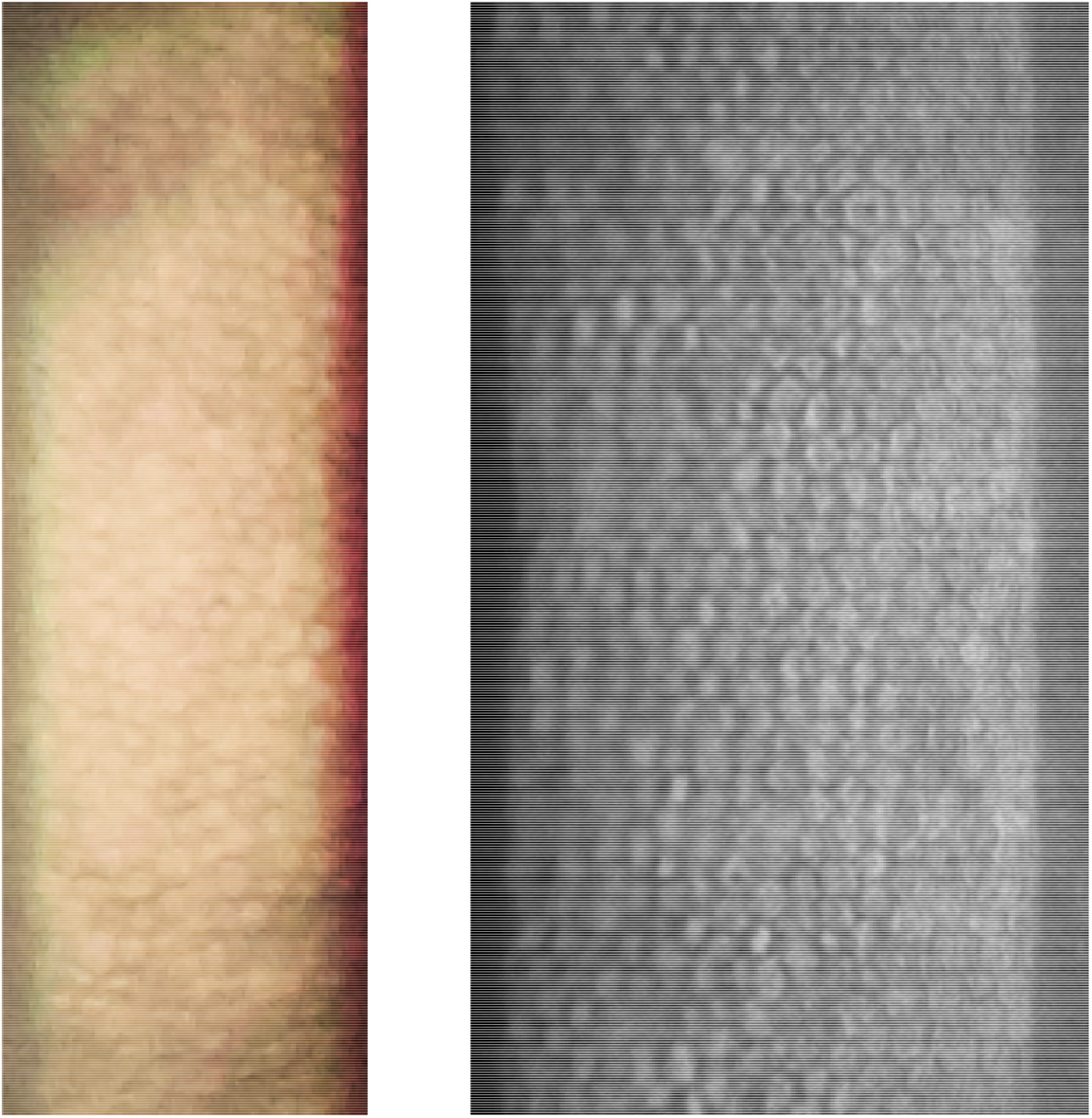
Comparison of Corneal Endothelial Images Captured by Smartphone-based Imaging (Left) and Captured by the Tomey Specular Microscope (Right). Both images show the characteristic honeycomb pattern of the corneal endothelium.

**Figure 3:**
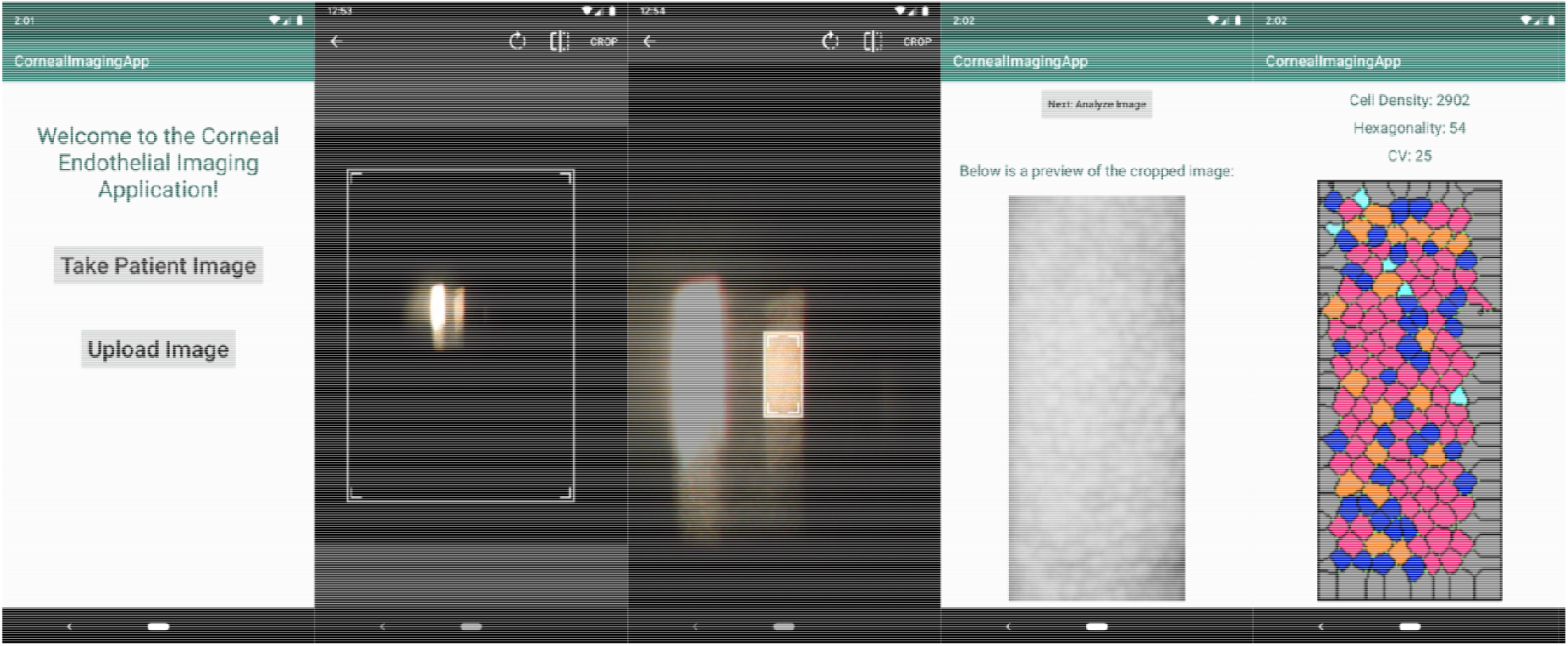
Image Acquisition and Analysis using Smartphone Application. a) User can upload or take image. b) Crop image and select sub-section for analysis. c) Preview of image to be processed. d) Results of analysis, including endothelial cell density (ECD), cell hexagonality (HEX), and cell variation (CV). Segmented image is displayed, with cyan cells being four-sided, blue cells being five-sided, pink cells being six-sided, orange cells being seven-sided, and white cells being eight-sided or more.

### Image Processing

In order to analyze the image and compute morphological parameters, we developed an image processing pipeline which consists of six steps: grayscale conversion, light normalization, image smoothing, KH segmentation, image binarization, thinning of border lines, artifact removal, and triple-point analysis. Each step of the process is displayed in Figure 4.

**Figure 4:**
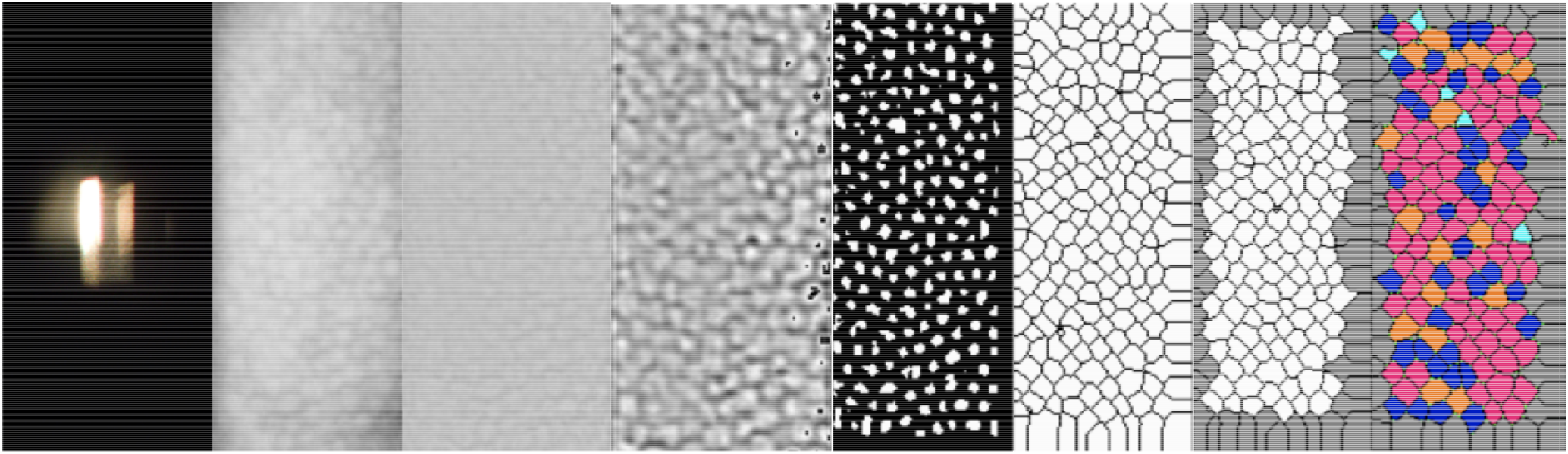
Image Analysis Processing Pipeline. Left to Right: Raw Image, Cropped Image, Light Normalization, Smoothing, KH Algorithm, Thinning, Artifact Removal, Triple-Point Analysis.

### Preprocessing

The image was first converted to grayscale, such that each pixel was stored as a value from 0 to 255. A smoothing method was applied to average the pixel values in a radius of 3 pixels to reduce image noise, remove artifacts, and amplify cell borders. Next, an algorithm was applied to normalize uneven illumination in the image. The technique adjusts pixel values in the image such that any given neighborhood of radius 16 pixels has approximately the same average brightness as the average brightness of the entire image.

### Segmentation

Once the two preprocessing steps were complete, the image underwent segmentation by a directional-filtering (KH) algorithm.^11^ Four masks were applied to the original image, and the resulting four images were merged to generate the final segmentation. However, occasionally cell nuclei still remained in the segmented images, disrupting further analysis. An artefact removal algorithm was applied to remove any objects less than 40 pixels in area, successfully filtering out both processing artefacts and nuclei.

A flood-based iterative thinning algorithm was used to thin all the cell borders in order to facilitate parameter calculation.^12^ A set of 3 by 3 masks were repeatedly applied to the segmented image, resulting in cell borders which are a single pixel wide.

### Parameter Calculation

In order to quantify cell shape, we used a triple point analysis technique.^13^ We identified the number of three-line intersections (triple points) that surround the cell, which is equivalent to the number of sides of a cell. Cell hexagonality was defined as the number of hexagonal cells divided by the total number of cells.

The cell density calculation was computed by counting the number of cells in the image and dividing by the actual size of the image (in mm^2^). The image size was inferred by the equation below, where *M*_s_ is the magnification of the slit-lamp microscope, *f*_C_ is the focal length of the OnePlus 7 Pro camera, *D*_v_ is the distance between the virtual image and the camera lens, *P*_a_ is the area of a single pixel on the OnePlus 7 Pro camera (in mm^2^), and *N* is the number of pixels in the selected image-processing area. The overall magnification (*M*_o_) was used to compute the size of each pixel, enabling calculation of endothelial cell density.

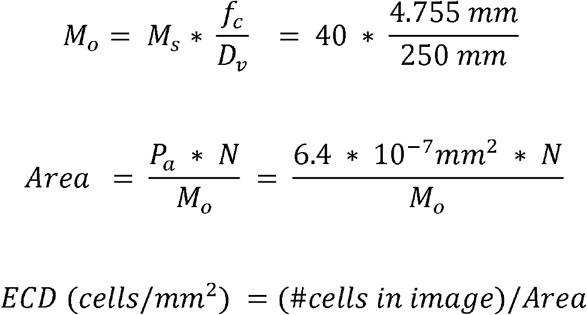

CV was computed by calculating the area of each cell and using the below equation, where µ represents the mean cell area (in pixels), represents the total number of cells, and *C*_i_ represents the size of a specific cell (in pixels).

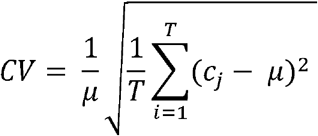

### Statistical Analysis

The data was analyzed using Python version 3.6 with packages scikit-learn and matplotlib. A paired Student’s t-test and Bland-Altman analysis was conducted.^14^ P-values < 0.05 were considered statistically significant.

Additionally, linear regression was performed on the Bland-Altman plots to assess proportional bias. The independent variable was the mean measurement, and the dependent variable was the difference between the specular and smartphone measurements. The null hypothesis was defined as the coefficient for the mean measurement being zero, and a two-sided Wald Test was used to compute the p-value.

## RESULTS

All 15 participants (30 eyes) in the study were healthy and did not have any known ocular diseases. The mean age of the participants was 29 ± 14 years, with a range from 19 to 72 years. The average number of cells analyzed by our smartphone algorithm was 185.56 ± 49.28 cells, and by the Tomey specular microscope was 239.86 ± 59.79 cells.

We compared the ECD, HEX, and CV computed by the Tomey specular microscope (-T) with those computed by the smartphone app (-S) by using paired two-sided t-testing. There was no significant difference in the mean ECD computed by the two devices. The mean ECD-T was 2799 ± 156 cells/mm^2^, and the mean ECD-S was 2779 ± 166 cells/mm^2^, with a mean difference of 20 cells/mm^2^ (p = 0.28). There was no significant difference in the mean HEX computed by the two devices. The mean HEX-T was 52 ± 6%, and the mean HEX-S was 53 ± 6%, with a mean difference of 1% (p = 0.50). There was a significant difference in the mean CV computed by the two devices. The mean CV-T was 34 ± 3% and the mean CV-S was 30 ± 3%, with a mean difference of 3.8% (p < 0.01).

Bland-Altman plots are a widely used method to assess concordance between two instruments, and plots for ECD, HEX, and CV are provided in Figure 5. The plots show that 60% of our ECD-S measurements differed by fewer than 100 cells/mm^2^ from the ECD-T. Similarly, 73% of HEX measurements had a smaller than 5% difference, and 96% of HEX measurements differed by less than 10%. Lastly, 70% of CV measurements had a smaller than 5% difference, and 97% of CV measurements differed by less than 10%. These data confirm that there is high concordance between ECD, HEX, and CV values as computed by the Tomey and our smartphone-based approach.

**Figure 5:**
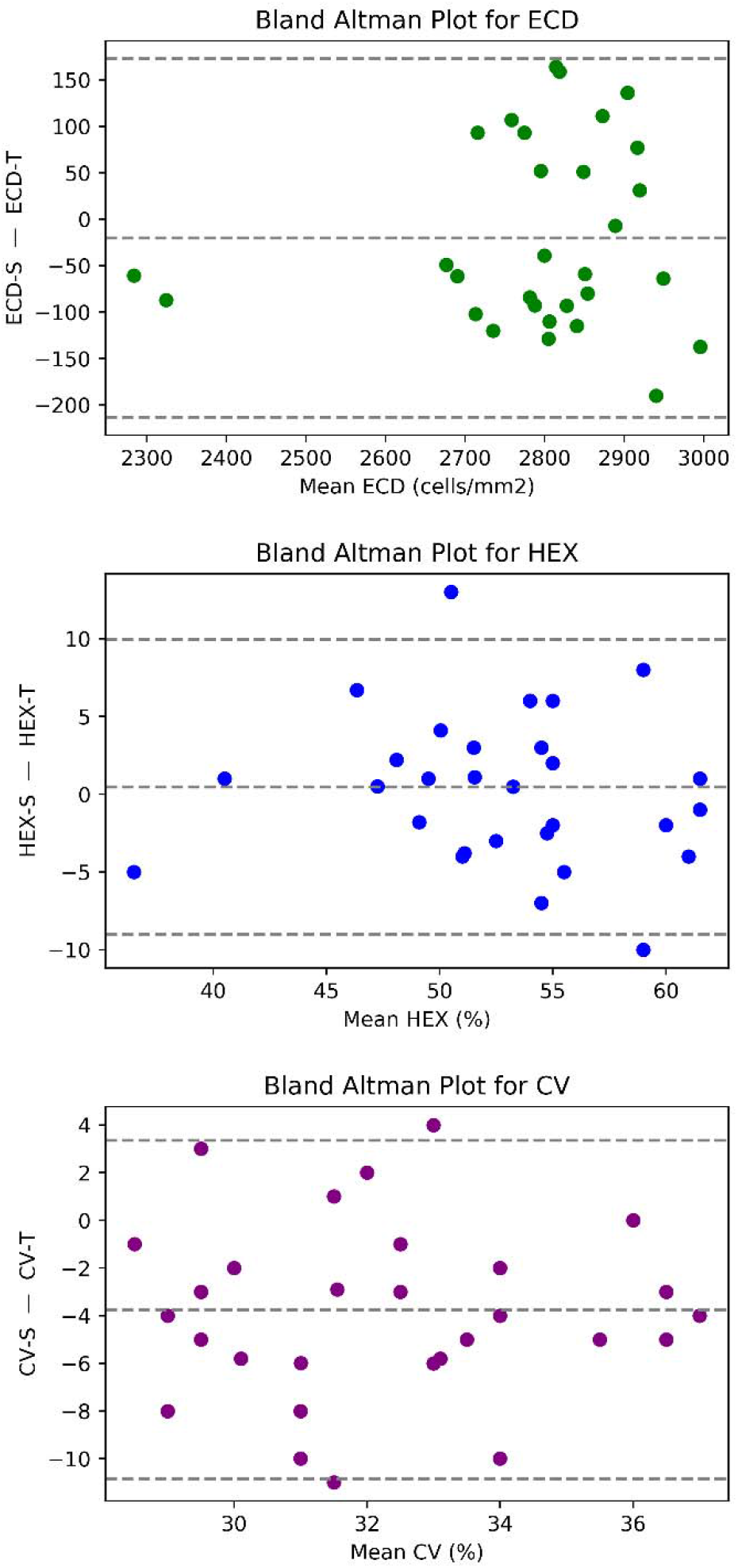
Bland-Altman Analysis comparing Smartphone-Based Imaging and Tomey Specular Microscope. The x-axis is the difference between the two measurements and the y-axis is the mean of the two measurements. The upper and lower dotted lines represent the 95% confidence interval. a) Comparison of Smartphone-Computed Endothelial Cell Density (ECD-S) and Tomey-Computed Endothelial Cell Density (ECD-T) b) Comparison of Smartphone-Computed Percentage of Hexagonal Cells (HEX-S) and Tomey-Computed Percentage of Hexagonal Cells (HEX-T) c) Comparison of Smartphone-Computed Cell Variation (CV-S) and Tomey-Computed Cell Variation (CV-T).

Linear regression was performed on the Bland-Altman plots to assess proportional bias. No proportional bias for ECD, HEX, and CV (p-values 0.58, 0.65, and 0.90, respectively) was found, indicating that the magnitude of the measurement is not correlated with the bias between the instruments.

## DISCUSSION

Our study demonstrates that smartphone-based imaging and analysis of the CE generates results comparable to those of traditional specular microscopy methods, providing a novel and low-cost tool for assessing CE function.

The mean ECD-T, HEX-T, and CV-T are very similar to those from previous studies which also imaged healthy subjects using the Tomey EM-4000 specular microscope.^15,16^ We found no significant difference between the ECD and HEX values computed by our smartphone-based algorithm and those provided by the Tomey specular microscope. Our Bland-Altman regression analysis demonstrated no proportional bias for all three measurements. We did find a statistically significant difference in the CV computed by the smartphone-based method and the Tomey specular microscope. However, previous studies have compared non-contact specular microscopes and have shown that there are statistically significant differences between device measurements of ECD, HEX and CV.^17,18^

Specifically, Gasser et al. compared the performance of the Topcon SP3000P and Konan Noncon Robo SP800 by imaging 34 healthy eyes and found that there was a statistically significant difference between the device measurements of ECD and CV. Similarly, Karaca et al. imaged 50 healthy eyes to compare the Nidek CEM-530 and Konan CellCheckXL and also found a statistically significant difference between device measurements of ECD and CV. Thus, the concordance we achieved we achieved between our smartphone-based method and the Tomey instrument is comparable to the concordance between clinically approved specular microscopes.

Previous studies have described smartphone-based endothelial imaging but had several limitations including a lack of automated analysis and a non-standardized imaging technique.^7^ Fliotsos et al. held a smartphone up by hand to the slit-lamp ocular and recorded a 3-minute long video of the specular reflection for each eye. This technique caused the patient to be subjected to bright light for an extended period of time and required an ophthalmologist to manually sort through each video and identify frames suitable for analysis. Our method of physically mounting the camera to the slit-lamp removes noise and artifacts generated by hand-movement. Further, we optimized the smartphone’s camera settings (including ISO and shutter speed) and standardized the slit-lamp’s beam. By keeping these factors the same between all patients, we were able to reliably and reproducibly image the endothelium of both eyes. The only factor that had to be adjusted in each session was the smartphone’s manual focus in order to compensate for the patient’s head movement. Furthermore, it took only 2-4 minutes to image and analyze both eyes on the smartphone device, which was similar to the speed of specular microscopy.

Additionally, all image analysis was performed in under 15 seconds on a mid-range Android smartphone, without the need for an internet connection or cloud computing services. Our image processing pipeline had robust performance across all images analyzed and was able to automatically compute clinically-relevant parameters (ECD, HEX, and CV) commonly used for monitoring endothelial health. To our knowledge, our work is the first to demonstrate that fully-automated analysis of smartphone-acquired corneal endothelium images is possible and generates results concordant with the analysis done by specular microscopes.

Our study illustrates the feasibility and accessibility of automated smartphone-based imaging and analysis of the CE. We leveraged a $500 smartphone and a $10 plastic attachment to image the CE at subcellular resolution. Commercial specular microscopes cost between $25,000 to $30,000 and are typically only available in tertiary-care facilities.^19^ Meanwhile, the slit-lamp is nearly ubiquitous in all ophthalmological clinics around the world, and over 90% percent of healthcare workers in many countries own smartphones.^20^ Moreover, many hospitals in developing countries conduct oureach eye-camps, where slit-lamps are brought into a rural field setting to assess patients.^21^ This technology would enable mobile, field-screening of endothelial health which is not possible through traditional means. Additionally, because of the app-guided nature of this technique, the screenings can be carried out by a technician or a community health worker, without requiring an ophthalmologist to be present. This would help make trained specialists available for more clinically urgent treatment roles.

The current limitations of this method include the requirement of a slit lamp with adequate magnification (32x or 40x) and the technical skill needed to obtain images. In the future, an auto-focus algorithm could be implemented to improve the ease-of-use of this technology. Furthermore, while we demonstrated high performance by using an image-processing pipeline with the KH algorithm to segment the smartphone images, additional image-processing methods could be explored and evaluated.

While our study demonstrates the promise of this method, it has a relatively small sample size of 30 eyes, which may limit the generalizability and statistical significance of the results. Future studies with a larger patient sample size, including patients with ocular diseases such as Fuchs endothelial corneal dystrophy (FECD), are required to further validate this technology. Preliminary data indicate that this technique can be used to image the CE, including guttae, in patients with FECD, as shown in Supplementary Figure 1. In addition, because the slit-lamp can be manually steered, our method could enable imaging of any central or peripheral region of the CE, rather than solely the 4-6 discrete regions imaged by commercial specular microscopes.

Overall, smartphone-based imaging is a promising low-cost technique that can assess the corneal endothelial status of healthy patients, with the potential to screen for endothelial disorders and identify patients at risk for corneal edema after intraocular surgery. Our study demonstrates that this method produces results concordant with those of a modern specular microscope, validating the performance of our novel image processing pipeline. The platform we developed is a fraction of the cost of current specular microscopes, is portable, and does not require an internet connection. This technology could enable regular endothelial screening in under-resourced communities, mitigating health disparities in eye care.

## Data Availability

Data will be made available upon reasonable request.

## ACKNOWLEDGEMENTS

We would like to acknowledge the participants of this study for their time. We thank Dr. GUMV Prasad and Dr. Venkatesh Lakkireddy from the Sri Kiran Institute of Ophthalmology for their assistance in imaging patients and thank the advisory board of the Global Alliance for Medical Innovation (GAMI) for their thoughtful feedback.

## CONFLICT OF INTEREST

The authors have no conflicts of interest or competing interests to report.

## FUNDING

The authors received no financial support when completing this study.

---

**Supplementary Figure 1:**
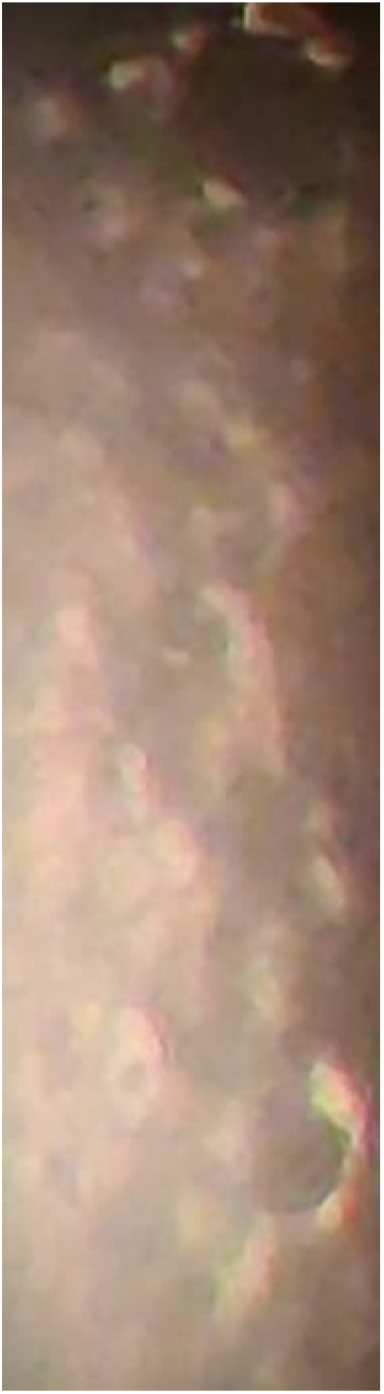
Smartphone-based Imaging of Corneal Endothelium of Patient with Fuchs Endothelial Dystrophy. Distinct guttae and excrescences are clearly visible.

## Notes

### Competing Interest Statement

The authors have declared no competing interest.

### Funding Statement

The authors did not receive any funding or compensation for this study.

### Author Declarations

This study was approved by the Institutional Review Board of the Sri Kiran Institute of Ophthalmology and was in compliance with the Declaration of Helsinki. Informed written consent was obtained from all participants.

## REFERENCES

1. Bourne WM. Biology of the corneal endothelium in health and disease. Eye. 2003. doi:10.1038/sj.eye.6700559

2. Musch DC, Niziol LM, Stein JD, Kamyar RM, Sugar A. Prevalence of corneal dystrophies in the United States: Estimates from claims data. Investigative Ophthalmology and Visual Science. 2011. doi:10.1167/iovs.11-7771

3. Edelstein SL, DeMatteo J, Stoeger CG, MacSai MS, Wang CH. Report of the Eye Bank Association of America Medical Review Subcommittee on Adverse Reactions Reported from 2007 to 2014. Cornea. 2016. doi:10.1097/ICO.0000000000000869

4. Ong Tone S, Jurkunas U. Imaging the Corneal Endothelium in Fuchs Corneal Endothelial Dystrophy. Seminars in Ophthalmology. 2019. doi:10.1080/08820538.2019.1632355

5. National Coverage Determination (NCD) for Endothelial Cell Photography (80.8). https://www.cms.gov/medicare-coverage-database/details/ncd-details.aspx?NCDId=213&ncdver=1&bc=AAAAQAAAAAAA&. Accessed November 25, 2020.

6. Toslak D, Thapa D, Erol MK, Chen Y, Yao X. Smartphone-based imaging of the corneal endothelium at sub-cellular resolution. Journal of Modern Optics. 2017. doi:10.1080/09500340.2016.1267815

7. Fliotsos MJ, Deljookorani S, Dzhaber D, Chandan S, Ighani M, Eghrari AO. Qualitative and Quantitative Analysis of the Corneal Endothelium With Smartphone Specular Microscopy. Cornea. February 2020. doi:10.1097/ICO.0000000000002277

8. Piorkowski A, Nurzynska K, Gronkowska-Serafin J, Selig B, Boldak C, Reska D. Influence of applied corneal endothelium image segmentation techniques on the clinical parameters. Computerized Medical Imaging and Graphics. 2017. doi:10.1016/j.compmedimag.2016.07.010

9. Daniel MC, Atzrodt L, Bucher F, et al. Automated segmentation of the corneal endothelium in a large set of ‘real-world’ specular microscopy images using the U-Net architecture. Scientific Reports. 2019. doi:10.1038/s41598-019-41034-2

10. Fabijańska A. Segmentation of corneal endothelium images using a U-Net-based convolutional neural network. Artificial Intelligence in Medicine. 2018. doi:10.1016/j.artmed.2018.04.004

11. Habrat K, Habrat M, Gronkow-Skaserafin J, Piórkowski A. Cell detection in corneal endothelial images using directional filters. In: Advances in Intelligent Systems and Computing. ; 2016. doi:10.1007/978-3-319-23814-2_14

12. Piórkowski A. Best-fit segmentation created using flood-based iterative thinning. In: Advances in Intelligent Systems and Computing. ; 2017. doi:10.1007/978-3-319-47274-4_7

13. Gronkowska-Serafin J, Piórkowski A. Corneal endothelial grid structure factor based on coefficient of variation of the cell sides lengths. In: Advances in Intelligent Systems and Computing. ; 2014. doi:10.1007/978-3-319-01622-1_2

14. Martin Bland J, Altman DG. Statistical Methods for Assessing Agreement Between Two Methods of Clinical Measurement. The Lancet. 1986. doi:10.1016/S0140-6736(86)90837-8

15. Kosekahya P, Ucgul Atilgan C, Atilgan KG, et al. Corneal Endothelial Morphology and Thickness Changes in Patients with Gout. Turkish journal of ophthalmology. 2019;49(4):178–182. doi:10.4274/tjo.galenos.2018.01947

16. Szalai E, Nemeth G, Berta A, Modis LJ. Evaluation of the corneal endothelium using noncontact and contact specular microscopy. Cornea. 2011;30(5):567–570. doi:10.1097/ico.0b013e3182000807

17. Gasser L, Reinhard T, Böhringer D. Comparison of corneal endothelial cell measurements by two non-contact specular microscopes. BMC Ophthalmology. 2015. doi:10.1186/s12886-015-0068-1

18. Karaca I, Yilmaz SG, Palamar M, Ates H. Comparison of central corneal thickness and endothelial cell measurements by Scheimpflug camera system and two noncontact specular microscopes. International Ophthalmology. 2018. doi:10.1007/s10792-017-0630-3

19. Bonnell AJ, Cymbor M. Under the Specular Microscope. Review of Optometry. 2012. doi:10.1016/S0747-5632(02)00016-X

20. Mobasheri MH, King D, Johnston M, Gautama S, Purkayastha S, Darzi A. The ownership and clinical use of smartphones by doctors and nurses in the UK: A multicentre survey study. BMJ Innovations. 2015. doi:10.1136/bmjinnov-2015-000062

21. Farooqui JH, Jorgenson R, Gomaa A. Mobilizing slit lamp to the field: A new affordable solution. In: Indian Journal of Ophthalmology. ; 2015. doi:10.4103/0301-4738.171972

